# Whole-Genome Sequencing is a Viable Replacement for Chromosomal Microarray and Fragile X PCR Testing

**DOI:** 10.1101/2025.05.24.25328260

**Authors:** Yueyao Gao, Sarah South, Colyn C Cain, Julie L Cox, Mark Fleharty, Benjamin A Hilton, Katie Larkin, Guang Li, David W Marsh, Victoria Popic, Reha M Toydemir, Sean Hofherr, Niall Lennon, Peter Bui

## Abstract

Developmental disabilities and congenital anomalies are common pediatric conditions that often require extensive genetic testing to determine an underlying cause. Traditionally, chromosomal microarrays (CMA) and Fragile X testing have served as first-tier diagnostics, but these tests are limited in scope and often necessitate follow-up sequencing assays. Whole Genome Sequencing (WGS) offers a single, comprehensive assay capable of detecting a broad spectrum of genetic variation, including single nucleotide variants (SNVs), insertions and deletions (INDELs), copy number variants (CNVs), structural variants (SVs), loss of heterozygosity (LOH), and tandem repeat alterations.

In this study, we evaluated whether WGS could replace CMA and serve as a more effective first-tier test. WGS achieved a 97.28% concordance with CMA for clinically relevant CNVs and LOH, while also offering more accurate breakpoint resolution and broader data point coverage. Notably, 4 out of 5 discordant cases (80%) were due to WGS providing more accurate breakpoint resolution. WGS covered over 97% of clinically relevant regions for CNV detection, compared to < 3% with CMA. To address the interpretive burden associated with the increased CNV calls, we implemented a cohort-based occurrence filter that successfully prioritized potential pathogenic events without sacrificing clinical sensitivity.

Additionally, we assessed the feasibility of Fragile X screening from WGS data using a custom PCR confirmation logic built on Expansion Hunter output. This approach accurately excluded normal-range alleles and flagged indeterminate or expanded alleles for follow-up PCR confirmation.

Our results support the use of WGS as a scalable and comprehensive diagnostic platform capable of consolidating multiple traditional assays. By streamlining workflows and enhancing clinical resolution, WGS offers a compelling alternative to the current diagnostic paradigm for patients with suspected genetic disorders.

## 1. Introduction

Developmental disabilities and congenital anomalies are relatively common pediatric conditions with lifelong implications that often result in long diagnostic odysseys for affected children and their families. In the United States, the estimated prevalence of any developmental disability among children between 2019 and 2021 ranged from 7.4% to 8.56%. Within the broader definition of developmental disability, intellectual disability was reported in approximately 2% of children, and autism spectrum disorder at 3%^1^. In Europe, the estimated prevalence of major congenital anomalies is approximately 2.4 cases per 100 live births according to the European Surveillance of Congenital Anomalies (EUROCAT)^2^.

Identifying an underlying etiology for developmental disabilities or congenital anomalies can inform medical management, guide prognosis, and improve access to appropriate interventions and support for the child and the family. Over the past decade, broader application of genetic testing has significantly expanded our understanding of the underlying causes of these conditions and further increased the diagnostic yield. This has led to improved patient outcomes, therapeutic options, and increased access to support networks for families^3^.

Chromosomal microarray (CMA) has long been recognized as a first-tier clinical genetic test for individuals with intellectual development disorders (IDD) and congenital anomalies^4,5,6^. CMA works by using millions of probes distributed across the genome to detect copy number variants (CNVs) and long regions of loss of heterozygosity (LOH). For example, the commonly used Affymetrix CytoScan HD platform contains over 2.67 million probes, including 1.9 million copy number probes and 750,000 SNP probes. This technology is optimized to detect clinically relevant CNVs in dosage-sensitive genes and LOHs that may indicate imprinting disorders or autosomal recessive conditions. However, its performance is constrained by the probe density and genomic coverage, leading to reduced sensitivity for smaller CNVs (< 50kb), limited resolution of breakpoints, and blind spots in off-target regions. Additionally, CMA can’t detect many other types of genetic variation, including single nucleotide variants (SNVs), insertions and deletions (INDELs), copy-neutral and complex structural variants (SVs), and trinucleotide repeats. These limitations can result in missed diagnoses, often necessitating follow-up with sequencing based assays or specialized tests like Fragile X analysis – either sequentially or in parallel. While parallel testing can expedite time to diagnosis, it often involves multiple billable tests performed at different laboratories with increased complexity and cost. In contrast, sequential or cascade testing can delay diagnosis and prolong the diagnostic odyssey. Although Whole Exome Sequencing (WES) and Whole Genome Sequencing (WGS) offer higher diagnostic yield, many patients still begin with CMA and Fragile X testing due to payors’ requirements. This stepwise approach often results in redundant testing, delayed diagnoses, and higher cumulative costs, both for families and the healthcare system.

Recent advances in next generation sequencing – including the reduced cost per sample, higher throughput, and the development of specialized analytical tools – have opened up new opportunities for WGS in clinical diagnostics. Current WGS analysis enables a comprehensive genomics assessment of a wide range of genetic variation, including SNVs, INDELs, SVs, CNVs, LOH, and tandem repeat alterations, all from a single assay.

In this study, we evaluate an alternative approach to traditional cascade or multi-assay testing by using WGS as a first-tier test. This approach enables both parallel and cascade testing strategies without requiring additional sample collections or multiple laboratory assays. By consolidating diverse test types into a unified WGS-based workflow in a single laboratory, this model reduces clinical molecular diagnostics laboratory complexity and enhances the overall diagnostic experience for patients.

We began by evaluating whether WGS could effectively replace CMA, which remains one of the most commonly ordered genetic tests for patients with suspected hereditary disorders^6,7^. We then extended our analysis to determine whether the same WGS data can reliably rule out Fragile X negative cases, aiming to triage negative cases, which would limit follow-up testing to a small subset of indeterminate, uncertain, and suspected positive cases requiring a confirmatory repeat primed PCR assay. Together, these two commonly ordered genetic tests – CMA and Fragile X – represent a critical benchmark for evaluating the clinical utility of WGS. Our findings show that WGS not only meets but exceeds the performance of these traditional assays, supporting its use as a comprehensive diagnostic platform capable of consolidating multiple laboratory assays into a single, efficient solution.

## 2. Methods

### 2.1 Sample Preparation and Sequencing

PCR-free whole genome sequencing (WGS) was performed on 142 DNA samples that were previously tested at Quest Diagnostics by Chromosomal Microarray (n = 103) or Fragile X repeat primed PCR (n = 39). This study was approved by an Institutional Review Board (WCG, IRB protocol identifier 20242498).

CMA samples were primarily whole blood-derived, collected in EDTA or Sodium Heparin tubes, with a few samples derived from buccal swabs. DNA was extracted using the QIAsymphony DSP DNA Mini Kit on the QIAsymphony SP instrument manufactured by Qiagen, which uses a magnetic-particle technology-based chemistry. After a lysis step, DNA was bound to magnetic particles, and the particle-bound DNA was washed and eluted in buffer.

Fragile X samples were extracted from whole blood collected in EDTA tubes using the MagNA Pure 96 DNA and Viral NA Small Volume Kit automated on the MagNA Pure 96 System from Roche, which utilizes magnetic glass particles (MGP) for purification. After a lysis step, DNA was bound to MagNA Pure Magnetic Glass Particles (MGP), and separated from the residual solution. Samples were washed on-bead to remove impurities, and DNA was eluted in buffer.

DNA was sent by Quest to Broad Clinical Labs for library preparation and sequencing. Initial concentration was measured by fluorescence-based dsDNA quantitation to ensure samples met the minimum input requirements for WGS. DNA was then mechanically sheared on the Covaris LE220-Plus and size-selected using AMPure XP beads. Libraries were prepared using the KAPA Hyper Prep PCR-free Kit, undergoing end repair, A-tailing, adapter ligation with indexed adapters, and qPCR-based normalization and pooling. Samples were sequenced to a mean depth of 30x on a 25B flowcell using NovaSeq X Plus, generating 150bp paired-end reads. The reads were aligned to the human reference genome GRCh37 using DRAGEN Secondary Analysis Platform v4.3.6 with default germline calling parameters.

### 2.2 WGS CNV Analysis

Aligned BAM files generated using DRAGEN v4.3.6 were further analyzed with an engineering build of the upcoming DRAGEN v4.4.1 release, which includes a cytogenetics module – a germline allele-specific copy number (ASCN) caller designed to produce CMA-equivalent results. This cytogenetics module incorporates features such as utilizing b-allele frequency in CNV calling, detection of LOH, smoothing of adjacent segments, mosaic event detection, and reporting of homozygosity index. To better tailor the calling to our evaluation needs, we applied three modifications to the default calling parameters: cnv-mosaic-fraction-cutoff 0.15, --cnv- interval-width 2,000, and --cnv-min-length-homozygosity-index 5,000,000.

### 2.3 Concordance Assessment with Clinically Reported Events from CMA

To evaluate the concordance of WGS-based CNV+LOH detection with clinically reported results from CMA, we evaluated 184 events from 99 de-identified samples, all previously tested on Affymetrix CytoScan HD microarray and analyzed using GRCh37. The CMA data were then analyzed using Chromosome Analysis Suite (ChAS) 4.5.0.34, with variant detection thresholds set at > 50 kb for DEL, > 200 kb for DUP, and > 5 Mb for LOH, with lower thresholds in regions of known clinical relevance. Further filtering using the Quest overlap map excluded variants with limited clinically relevant CNVs.

The remaining CNVs were then classified into one of five categories: pathogenic, likely pathogenic, variant of uncertain significance (VUS), likely benign, or benign. These categories are derived from the 2019 ACMG guidelines for interpretation and reporting of constitutional copy number variants^8^. Only pathogenic, likely pathogenic, and qualifying VUS (i.e., > 200 kb for DEL and > 500 kb for DUP) were reported. LOH regions were reported if they met predefined size and context criteria (e.g., 2% of the genome, terminal LOH > 5 Mb, or interstitial ROH size threshold based on chromosome imprinting status).

We then compiled the clinically reported events from each sample into a VCF file and compared them against the WGS-based calls from the ASCN module using the SV benchmarking tool “truvari”9. In this analysis, we considered a WGS call concordant with a CMA-reported event if it matched its event type and showed at least 70% reciprocal overlap and 70% size similarity (default setting in truvari). We did not apply a maximum breakpoint distance criteria, as large events (>10Mb) often have imprecise breakpoints in CMA due to probe design limitations. In such cases, the absolute distance between breakpoints across the two platforms can be substantial, making it difficult to define a consistent and biologically meaningful threshold.

### 2.4 CMA-WGS CNV Point Coverage Analysis

We assessed the genomic coverage available for CNV detection from two platforms: WGS and CMA. For WGS, the CNV data coverage BED file was generated by subtracting the CNV detection excluded regions ( *cnv.excluded_interval.bed.gz, included in DRAGEN CNV output) from the GRCh37 reference genome using “bedtools subtract”. For CMA, we used the probe coverage BED file from the CytoScan HD Array Annotation NA32, available as part of the software library files.

To quantify genome-wide coverage, “bedtools coverage” was used to calculate the proportion of bases covered by each platform across the genome using GRCh37 genome-wide BED files. Clinically relevant dosage-sensitive regions were extracted from the ClinGen Dosage Sensitivity Map^10^ by selecting only regions with sufficient evidence for haploinsufficiency (HI) or triplosensitivity (TS). We then applied “bedtools intersect” to identify overlaps between the CNV coverage regions and ClinGen dosage-sensitive intervals. Finally, “bedtools coverage” was again used to quantify base-level coverage in these clinically relevant regions, stratified by chromosome.

### 2.5 Cohort-based Occurrence Filter

We designed a cohort-based occurrence filtering scheme for DRAGEN ASCN calls to identify and exclude likely benign or artifactual CNVs. This filter was implemented using BCFtools, Truvari, and a custom Python script. First, CNV calls from a cohort of 103 samples were aggregated by ID using bcftools merge. The horizontally merged multi-sample VCF was then processed with Truvari collapse to merge overlapping variants based on the following criteria: ≥ 70% size similarity, ≥ 70% reciprocal overlap, and breakpoint proximity within 2kb. Next, we used “Truvari Anno gtcnt” to annotate each merged record with its occurrence count across the cohort, generating a consolidated cohort.cnv.vcf, in which each variant represents a distinct event with a corresponding occurrence in the cohort.

For each new sample, the ASCN output VCF was benchmarked against the cohort.cnv.vcf using the same matching threshold. Variants observed in more than five individuals in the cohort were considered recurrent and removed from the final WGS with filter output to reduce interpretation burden for variant analysts.

### 2.6 Visualization of CNVs Evidence from WGS and CMA

To facilitate visual validation of CNV and LOH calls, we combined the tracks from WGS and CMA. The details of each track are described in Table 1.

**Table 1.** Overview of Tracks Used for CNV and LOH visualization in WGS and CMA Data.

| Platform | Track | Description |
| --- | --- | --- |
| WGS | CNV excluded | Regions marked as KMER-NONUNIQ, which are unsuitable for CNV calling due to low mappability. |
|  | Target read counts | Raw read counts per 2kb target interval. |
|  | Normalized target read counts | Log2-normalized read count per 2kb target interval. |
|  | Segmentation of target read count | Segmentation of normalized target read count. |
|  | BAF | B-allele frequency of common SNPs (> 10% allele frequency from the 1k genome project) |
|  | Segmentation of BAF | Segmentation of b-allele loci |
|  | CNV + LOH Call | Indicator for regions containing CNV or LOH |
| CMA | CNV Call | Indicator for regions containing CNV |
|  | LOH Call | Indicator for regions containing LOH |
|  | Log2 Ratio | Log<sub>2</sub> ratio of normalized sample intensity vs. reference, adjusted for noise. |
|  | BAF | B-allele frequency of SNP probes |

### 2.7 Fragile X Rule Out with Expansion Hunter

A total of 39 samples with known FMR1 CGG repeat sizes, previously determined by repeat primed PCR, were selected for evaluation. The cohort included 16 normal (< 40 repeats), 12 gray zone (41-54 repeats), 4 permutation (55-199 repeats), and 7 full expansion cases (> 200 repeats). Repeat sizing from WGS was performed using Expansion Hunter^11^ which is integrated into the DRAGEN Secondary Analysis Platform v4.3.6.

To minimize false negatives in Fragile X screening, we implemented a confirmation logic based on Expansion Hunter output. Samples with low-confidence or borderline repeat size were flagged for PCR confirmation, while clearly negative results could be confidently excluded by WGS alone. Classification decisions were guided by three attributes from Expansion Hunter output: ADSP (the number of spanning reads that fully cover the CGG repeat region), REPCI (repeats confidence interval), REPCN (the estimated repeat copy number).

The confirmation logic was designed to account for the sex of the participant, as the number of FMR1 alleles differs between males and females. Assessing the minimum number of spanning reads (minADSP) across alleles helps identify cases where one or both alleles may be poorly supported, increasing the risk of a false negative classification. Similarly, the maximum width of the repeat confidence interval (maxREPCI delta) captures the uncertainty in repeat size estimation, which is especially relevant when allelic resolution is limited due to the nature of short-read sequencing.

When minADSP < 2 and maxREPCI delta ≥ 2, the result was considered unconfirmed by WGS and flagged for PCR confirmation due to insufficient spanning evidence and high sizing uncertainty. In rare cases where minADSP < 2 and maxREPCI delta < 2, REPCN < 35 were interpreted as negative, whereas REPCN ≥ 35 triggered PCR confirmation.

For samples with stronger spanning support (minADSP≥ 2), high confidence was assumed. In these cases, REPCN < 40 were confidently classified as negative, while those ≥ 40 will prompt PCR confirmation testing, though the likelihood of expansion was low.

## 3. Results

### 3.1 Performance concordance between CMA and WGS

We assessed the concordance between CMA and WGS using 99 de-identified samples across three runs, comprising a total of 184 reported events. These included 59 Deletions (DELs) and 47 Duplications (DUPs), ranging in size from 55kb to 39 Mb, and classified as pathogenic/likely pathogenic (42%) or variant of unknown significance (51%). Additionally, 78 loss of heterozygosity (LOH) regions from eight samples, including one full chromosome aneuploidy, were evaluated.

Among these, 57 of 59 DELs (96.61%, 95% CI: 88.45 - 99.07) and 46 of 47 DUPs (97.87%, 95% CI: 88.89 - 99.62) showed equivalent results between CMA and WGS, defined in Methods 2.3 (Table 2). Similarly, 76 of 78 (97.44%, 95%CI: 91.12-99.29) LOH regions were concordant between the two platforms. Two sex-chromosome events from one sample were initially missed by WGS but were later found upon manual review to represent the same events as CMA, though they were presented differently by the respective platforms (Supplemental Note 1). Another missed call involving a DUP was also an LOH; WGS annotated it as GAINLOH in the VCF, while CMA reported at DUP. This discrepancy caused Truvari to label it as False Negative, though it was ultimately a representational difference. Additionally, two CMA-reported CNV events were split into a few subcalls by WGS. Manual inspection showed that Affymetrix made similar subcalls, which were manually merged during variant interpretation by Quest (Supplemental Note 2).

**Table 2.** Summary of concordance between CMA and WGS for solves identified by CMA.

| Event Type | Size | CMA Detected Solves | Recovered by cWGS | Concordance(%) |
| --- | --- | --- | --- | --- |
| DEL | 50kb-100kb | 5 | 5 | 100.00 |
|  | 100kb-500kb | 17 | 16 | 94.12 |
|  | 500kb-1Mb | 7 | 7 | 100.00 |
|  | 1Mb-10Mb | 22 | 21 | 95.45 |
|  | 10Mb-25Mb | 4 | 4 | 100.00 |
|  | 25Mb+ | 4 | 4 | 100.00 |
|  | **Overall** | **59** | **57** | **96.61**<br>**(95% CI: 88.46-99.07)** |
| DUP | 100kb-500kb | 4 | 4 | 100.00 |
|  | 500kb-1Mb | 25 | 25 | 100.00 |
|  | 1Mb-10Mb | 16 | 15 | 93.75 |
|  | 10Mb-25Mb | 1 | 1 | 100.00 |
|  | 25Mb+ | 1 | 1 | 100.00 |
|  | **Overall** | **47** | **46** | **97.87**<br>**(95% CI: 88.89-99.62)** |
| LOH | 5Mb-10Mb | 30 | 29 | 96.67 |
|  | 10Mb-25Mb | 36 | 35 | 97.22 |
|  | 25Mb+ | 12 | 12 | 100.00 |
|  | **Overall** | **78** | **76** | **97.44**<br>**(95% CI: 91.12-99.29)** |
| All |  | 184 | 179 | 97.28<br>(95% CI: 93.80-98.83) |

Figure 1 shows the concordance between CMA and WGS prior to manual inspection. Across all 181 events, the average reciprocal overlap percentage is 95.91%, with a 95% confidence interval of 94.64% to 97.19%. This indicates that most CMA-reported events show strong concordance with WGS in terms of variant breakpoints.

**Figure 1:**
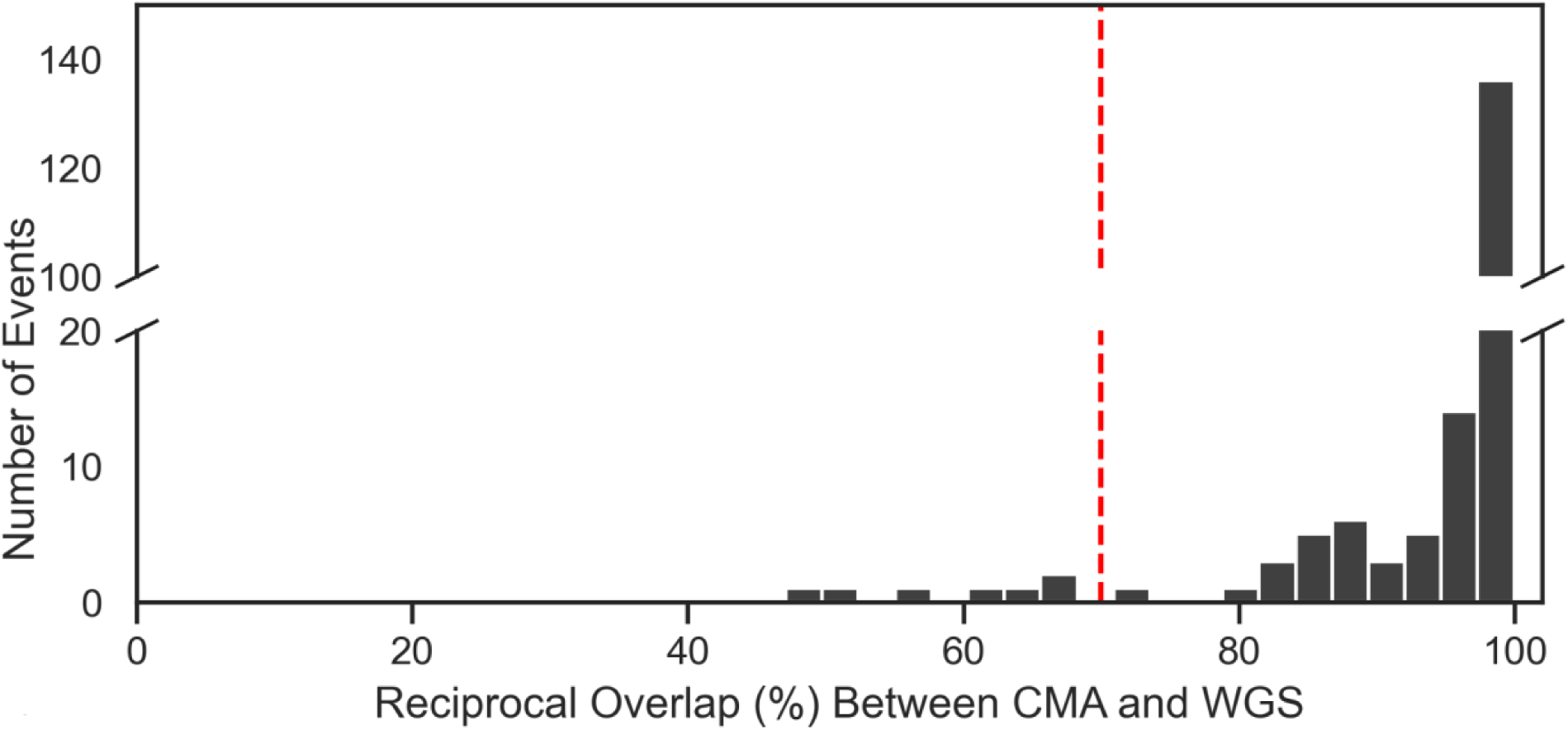
Reciprocal overlap percentage between CMA and WGS for clinical reported events prior to manual inspection. Histogram displaying the distribution of reciprocal overlap percentages between CMA-reported events and WGS-detected CNVs and LOH across 181 events. The x-axis represents the reciprocal overlap percentage, and the y-axis indicates the number of events within each overlap range.

### 3.2 Evaluating Discordant CNV Calls between Two Platforms

To investigate differences in CNV detection between CMA and WGS, we manually reviewed the five events that did not meet the 70% reciprocal overlap matching threshold. These represent the only instances of suboptimal concordance (47.12% - 68.26% overlap) among the 184 evaluated events. A summary of these discordant cases is provided in Table 2. Notably, in four of the five cases, WGS demonstrated higher resolution and more accurate breakpoint refinement than CMA, largely due to the insufficient CMA probe coverage.

#### Case 1: 1.9Mb Pathogenic DEL on 1q21.1q21.2

A 1.9Mb heterozygous copy number loss on chromosome 1q21.1q21.2 was detected by CMA, consistent with a well-characterized 1q21.1 microdeletion syndrome^12,13,14,15^. As shown in Figure 2, WGS identified two heterozygous DELs in the same region, measuring 902.3kb and 97.6kb, separated by a 334.1kb gap. Additionally, the CMA call extended 390.2kb further on the left and 190.8kb on the right side, resulting in a total discrepancy of approximately 860kb between the two platforms.

**Figure 2:**
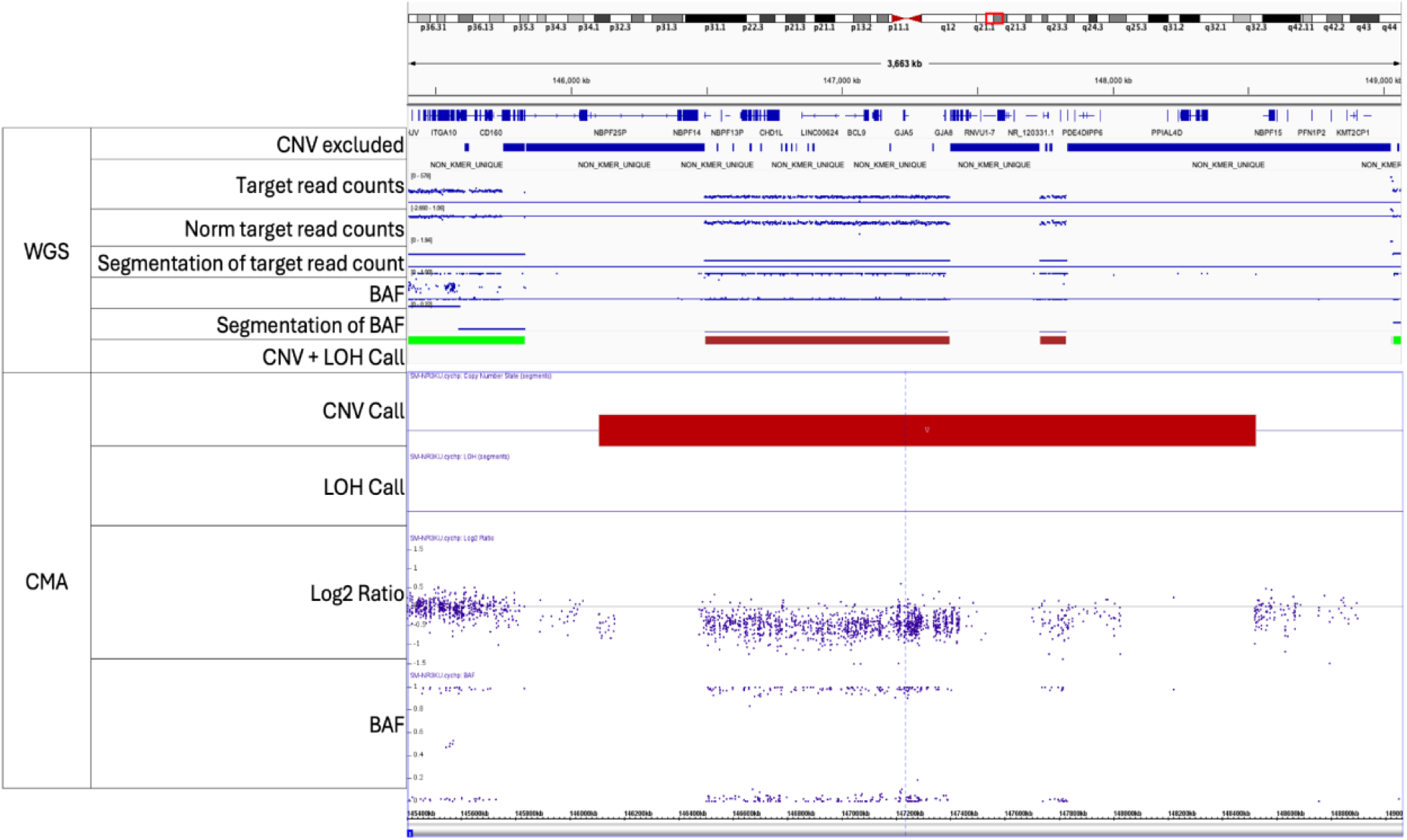
Integrated visualization of Case 1 using IGV (WGS) and Affymetrix (CMA). Tracks descriptions are provided in Methods section 2.5.

This discrepancy is primarily caused by the NON-KMER UNIQUE CNV excluded region, where WGS sequencing depth is considered unreliable for CNV calling by DRAGEN. Notably, the 334.1kb gap between the two WGS CNV calls contains very few CMA probes, indicating that the data points in this region are less sufficiently compared to the surrounding regions. Similarly, the flanking regions included in the CMA call but not captured by WGS also have sparse probe coverage. While DRAGEN’s ASCN caller performs smoothing to merge adjacent segments with a similar normalized copy number ratio, the size of the NON-KMER UNIQUE region in this case exceeds the threshold for consolidation. Of the 184 evaluated events, this is the only instance where WGS failed to merge CNV segments into a single call with ≥ 70% reciprocal overlap with the corresponding CMA event.

#### Case 2: 312kb Pathogenic DEL on 15q11.2

A 312kb heterozygous DEL on 15q11.2 (chr15:22770422-23082328), encompassing the NIPA1 and NIPA2 genes, was detected by CMA. This copy number variant has been previously associated with highly variable clinical phenotypes and was reported in this case as an unrelated pathogenic CNV in a patient with seizures. WGS identified a larger heterozygous DEL at chr15:22751964-23223781, extending approximately 160kb downstream of the CMA reported interval.

This discrepancy is largely attributable to limited CMA probe coverage at the downstream end of the WGS call, as shown in Figure 3. Similarly, DRAGEN also excluded a region of 121.6kb at the downstream boundary due to insufficient mappable content. However, WGS captured additional informative data points beyond the excluded region, enabling the detection of another segment of DEL with a similar copy number ratio. Furthermore, the DRAGEN ASCN caller merged the two adjacent DEL calls due to their close proximity and consistent signal. Population SNP data within this extended region appeared exclusively as either homozygous alternate or homozygous reference in the WGS, supporting the validity of the smoothed DEL call. This case demonstrates the advantage of WGS in capturing extended CNV signals beyond the resolution limits imposed by CMA probe coverage.

**Figure 3:**
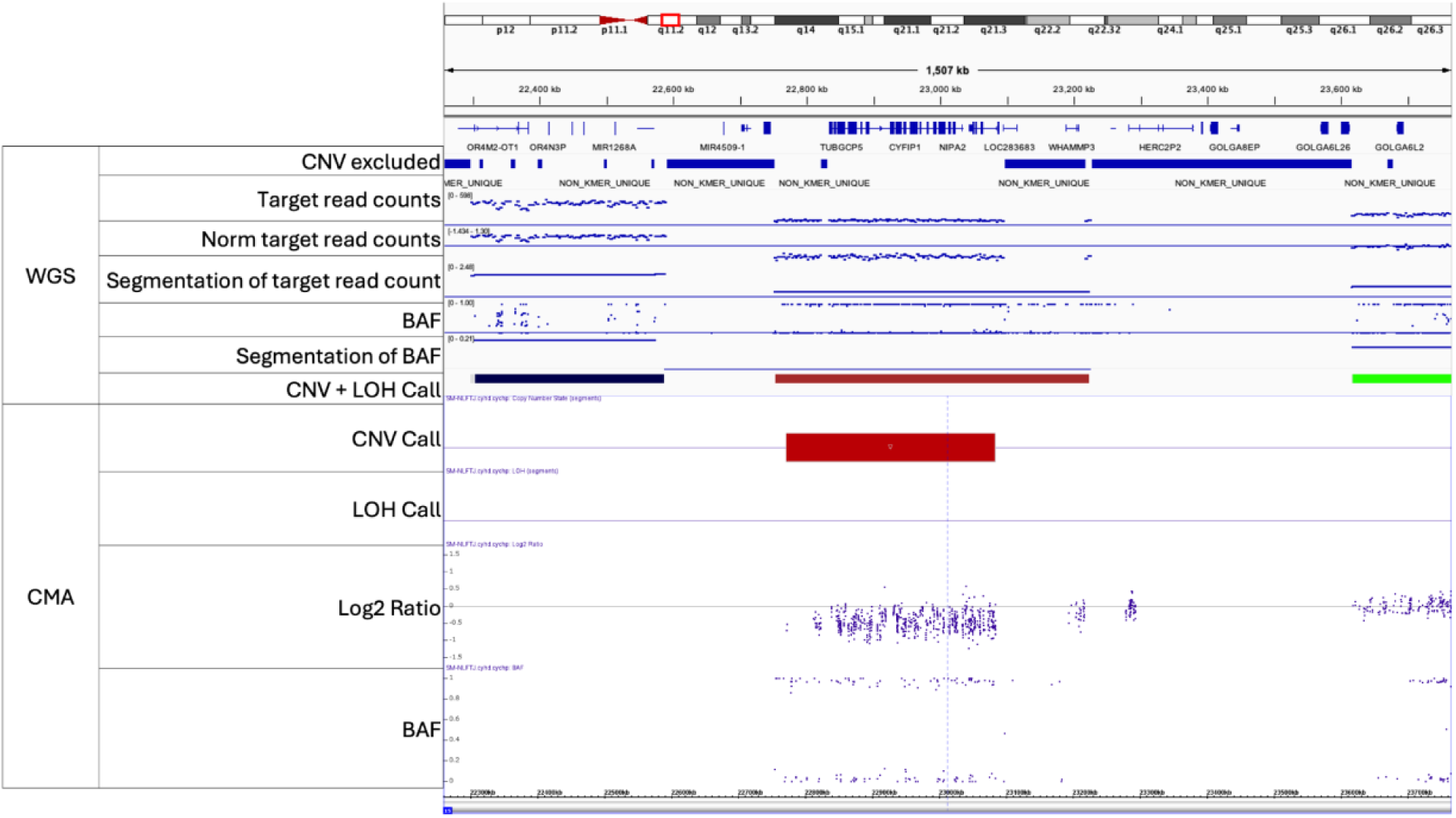
Integrated visualization of Case 2 using IGV (WGS) and Affymetrix (CMA). Tracks descriptions are provided in Methods section 2.5.

#### Case 3: 1.1Mb DUP on 22q11.21

A 1.1Mb copy number gain (CN3) on 22q11.21 (chr22:20728959-21800797) was detected by CMA and classified as a VUS. This CNV was part of a broader diagnosis of Turner syndrome, which also included a pathogenic complex structural abnormality on chrX. WGS identified a 731.7kb CN3 DUP that was 336.3kb shorter on the right end. As shown in Figure 4, there are very few CMA probes supporting the region not captured by WGS. At the right breakpoint, WGS data shows depth and BAF consistent with CN2, rather than a CN3 gain. This suggests that WGS may have provided a more accurate breakpoint on the right side, enabled by its better resolution and more continuous data coverage.

**Figure 4:**
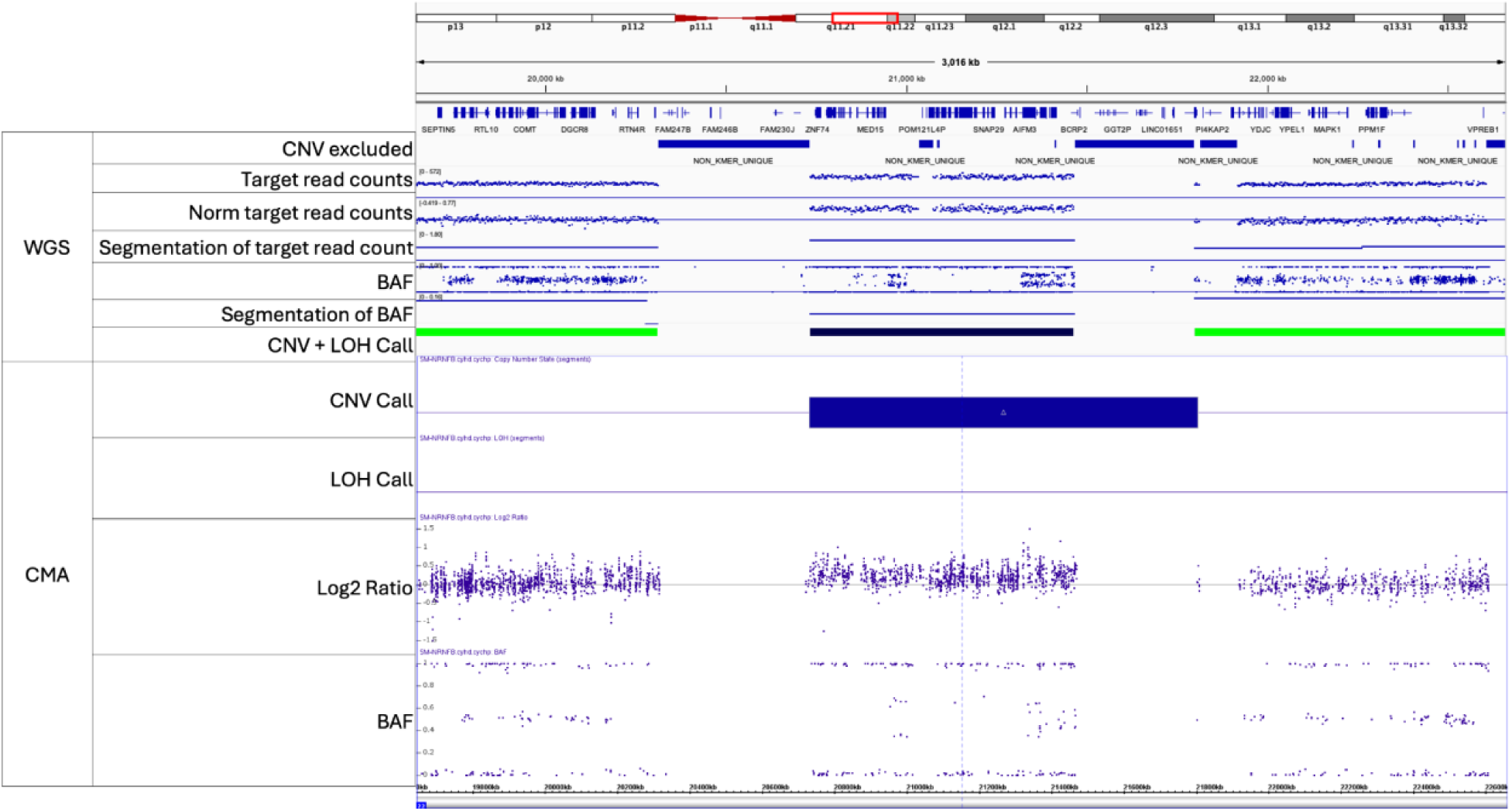
Integrated visualization of Case 3 using IGV (WGS) and Affymetrix (CMA). Tracks descriptions are provided in Methods section 2.5.

#### Case 4: 15.8Mb LOH

A 15Mb LOH on 1q21.1q23.2 was detected by CMA as part of multiple reported homozygous regions in an individual. WGS identified two LOH segments in this region, measuring 9.9Mb and 2Mb, respectively. As shown in Figure 5, the region not flagged as LOH by WGS displays clear evidence of heterozygosity in the BAF track. In contrast, CMA failed to capture the variability of this region, likely due to insufficient probe coverage. This case highlights how WGS, with its higher density of informative data points, enables more accurate LOH detection compared to CMA.

**Figure 5:**
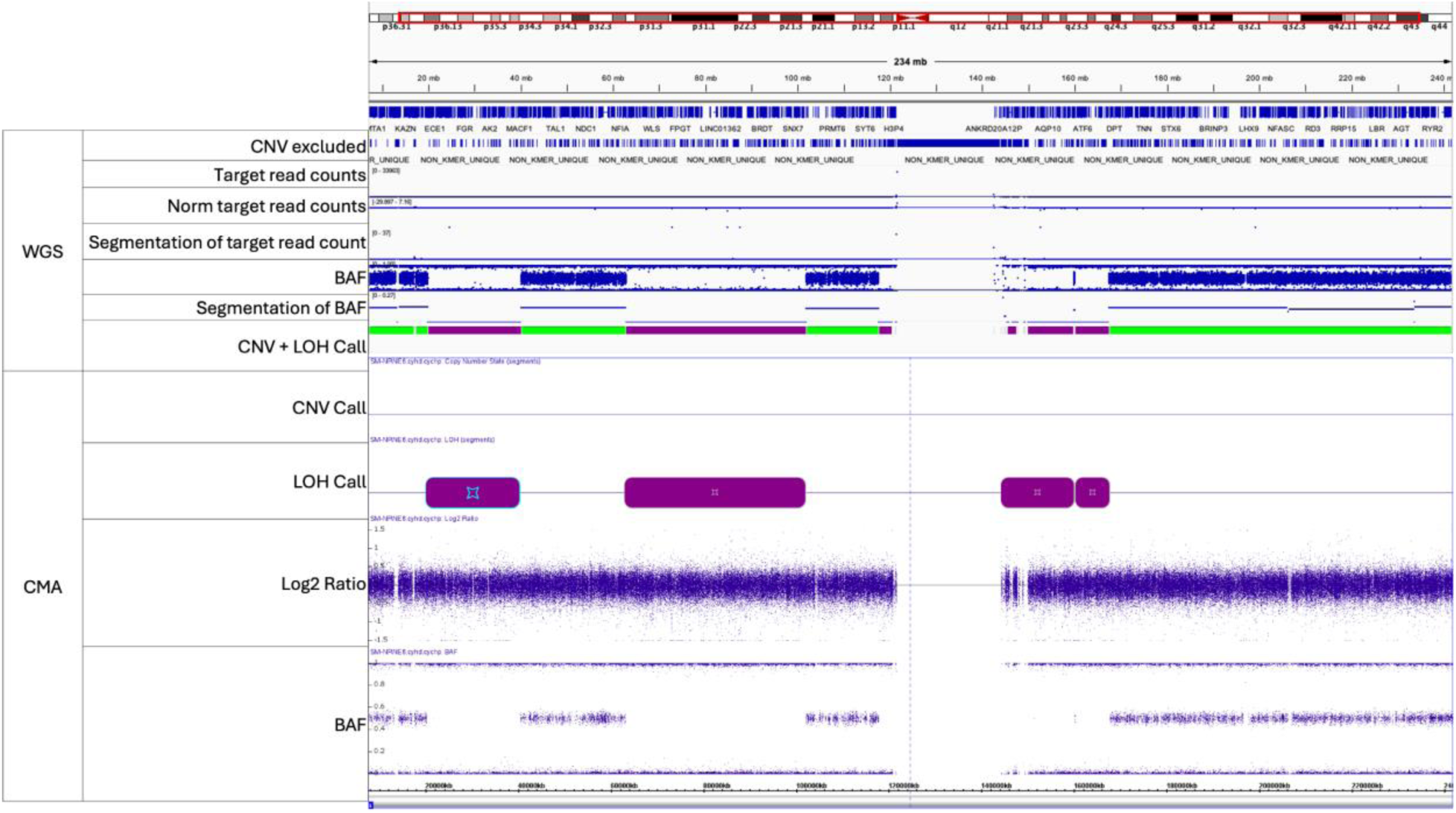
Integrated visualization of Case 4 using IGV (WGS) and Affymetrix (CMA). Tracks descriptions are provided in Methods section 2.5.

#### Case 5: 7Mb LOH

A 7Mb LOH on 16p11.2p11.1 was detected by CMA as part of multiple reported homozygous regions in an individual. In comparison, WGS identified two smaller LOH segments within this region, measuring 1Mb and 3.9Mb, resulting in a 2.1Mb discordant region between the two platforms. As shown in Figure 6, the region not flagged as LOH by WGS displays clear heterozygosity in the BAF track, suggesting real variability that CMA failed to detect – likely due to sparse probe coverage across the discordant interval. Similar to Case 4, this example illustrates WGS’s superior resolution for LOH detection, which may be misclassified by array based methods.

**Figure 6:**
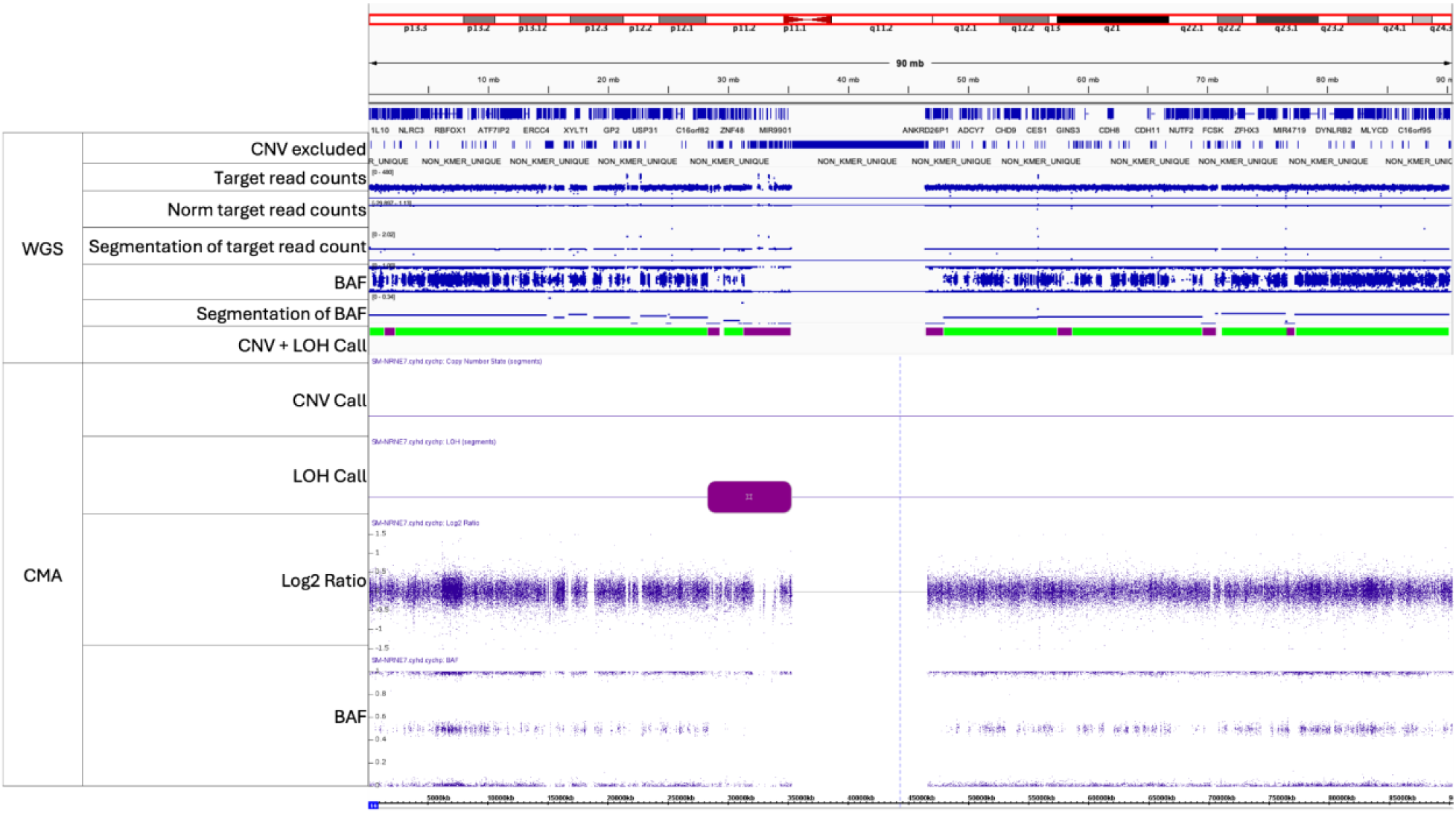
Integrated visualization of Case 5 using IGV (WGS) and Affymetrix (CMA). Tracks descriptions are provided in Methods section 2.5.

Together, these case reviews highlight how the higher resolution of WGS can provide more precise breakpoint determination compared to CMA. The improved granularity of WGS, enabled by continuous genomic coverage and dense variant data, provides a more precise and nuanced view of CNV and LOH events. In the following section, we broaden our focus from individual cases to a genome-wide, systematic evaluation of CNV calling coverage difference between the two platforms.

### 3.3 CNV Calling Coverage Difference CMA and WGS

To compare the resolution of data available for CNV detection, we evaluated the genomic coverage between two platforms across the whole genome as well as a set of dosage-sensitive regions from ClinGen. As shown in Figure 7, WGS consistently provides substantially higher data point coverage than CMA. Across the genome, the average data point coverage for CNV calling with CMA is 1.33% (95% CI: 1.22-1.43), compared to 83.55% with WGS (95% CI: 76.59- 90.52). This disparity is further highlighted when focusing on regions with sufficient evidence for haploinsufficiency (HI) and triplosensitivity (TS) according to the ClinGen Dosage Sensitivity map. In these clinically relevant regions, CMA achieves an average coverage of 2.29% (95% CI: 1.90-2.49), while WGS achieves 97.38% (CI: 96.24-98.53). These findings demonstrate that WGS offers significantly more granular and comprehensive representation of the genome for CNV calling, which is critical for accurate breakpoint determination and characterization of CNVs.

**Figure 7:**
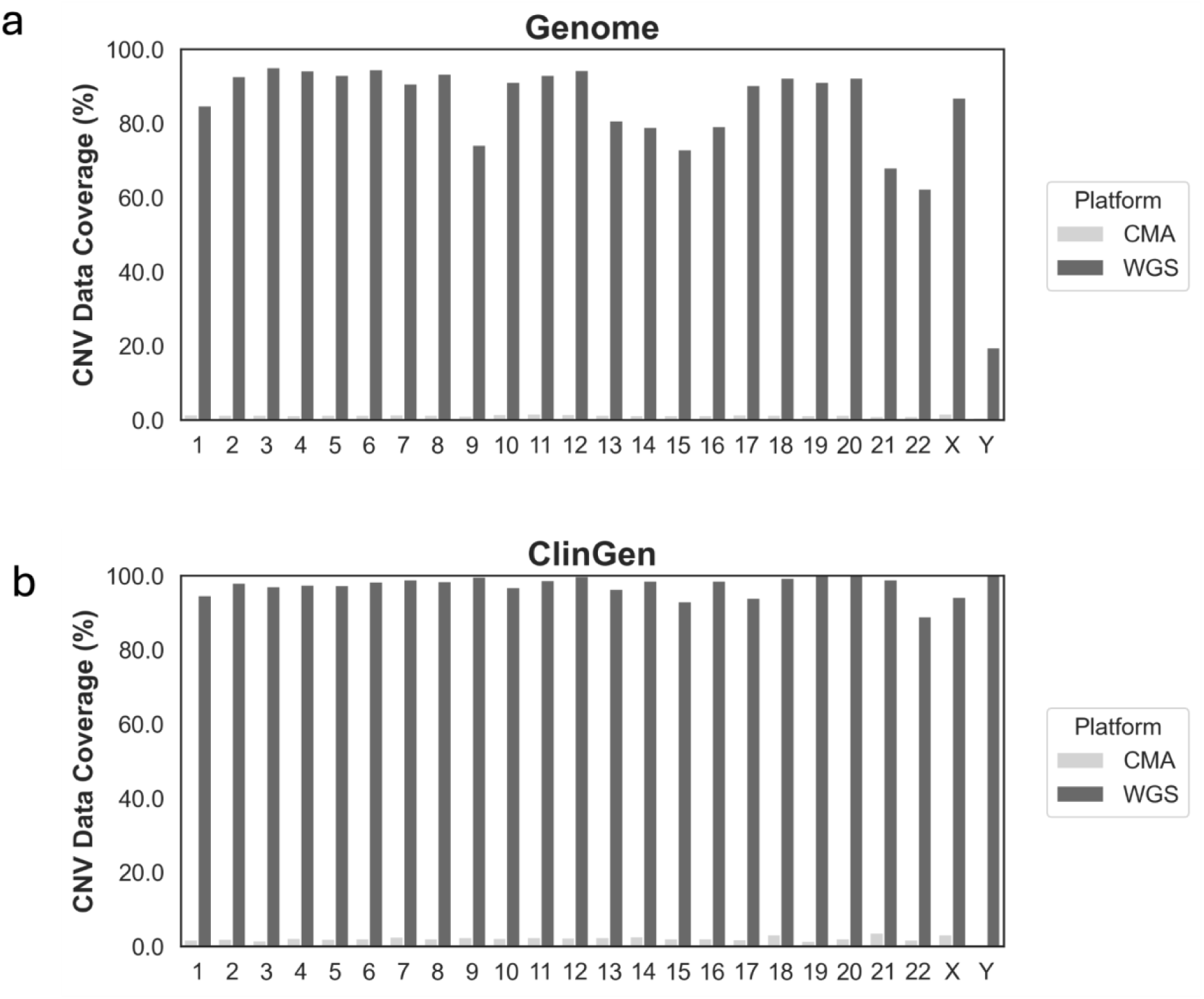
Comparison of CNV data point coverage across platforms and genomic regions. Bar plots show the percentage of genomic regions covered CNV data points from CMA and WGS across chromosomes. The left panel represents the whole genome coverage, while the left panel focuses on ClinGen dosage-sensitivity region (sufficient evidence for haploinsufficiency or triplosensitivity). The x-axis indicates chromosomes, and the y-axis indicates the percentage of bases with CNV-supporting data points. WGS demonstrates substantially higher coverage than CMA in both genome-wide and ClinGen-defined dosage sensitive regions.

### 3.4 Number of Events for Interpretation from CMA and WGS

A common concern with the higher resolution and broader data coverage of WGS is the increased number of CNV calls requiring interpretation. Even with the DRAGEN ASCN caller, which is designed to approximate CMA-level findings, the raw VCF typically contains over 100 PASS-filtered events per case. While many of these calls are likely biologically valid, they substantially increase the manual workflow for variant analysts during clinical interpretation.

To address this, we developed a customized filtering strategy to retain clinically relevant events while minimizing false positives and common population variants. First, we applied the same size threshold commonly used in the CMA workflow: DEL > 50kb, DUP > 200kb, LOH > 5Mb. To avoid redundancy, overlapping records with different resolutions (RES) were collapsed to retain only the highest-RES call. Each WGS record was then compared against a 103-sample internal WGS cohort from Quest. Any event observed in more than five individuals was considered non-clinically significant and excluded. For an average case, these steps removed ∼95% of events yielding an average of eight (95% CI: 6.63-9.25) – comparable to typical CMA-based review workloads.

Importantly, all clinically reported CNVs and LOH were retained post-filtering except for case5 in 3.2. In that instance, WGS accurately identified two LOH segments (3.9Mb and 1Mb), but the 5Mb size threshold excluded them. Since the clinical report described this sample as having no copy number variants identified; multiple regions of homozygosity detected, the diagnostic sensitivity was not compromised.

Figure 8 summarized the number of calls per case across four categories: raw CMA calls, CMA calls after applying an internal filter (which includes common events and CMA specific artifacts), raw WGS calls, and WGS calls after applying a cohort-based recurrent filter. The comparison is based on the 99 samples used in this evaluation. For consistency, both raw CMA and raw WGS results were subject to the same size threshold described above. As expected, WGS initially yields more events than CMA when only equivalent size filters are applied. However, once the cohort-based recurrence filter is applied, the final number of WGS events per case aligns closely with the filtered CMA results.

**Figure 8:**
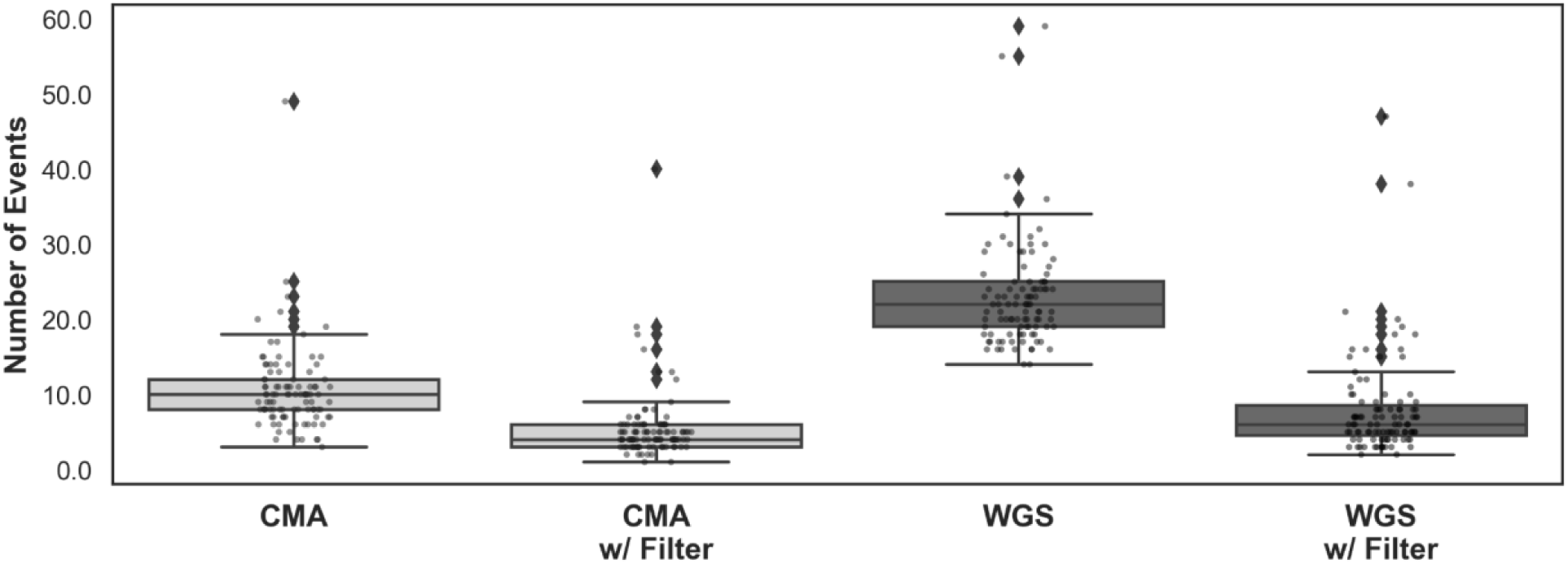
Comparison of the total number of calls between CMA and WGS with and without filter. The number of calls (CNV+LOH) per sample across 99 cases under four conditions: unfiltered CMA, filtered CMA, unfiltered WGS, and filtered WGS. A consistent CNV size threshold was applied for comparability. The box plots display the interquartile range (IQR, the middle 50% of the data), with whiskers extending to 1.5x of the IQR. Points beyond the whiskers representing outliners. An overlaid strip plot displays all individual data points within each group.

Overall, this demonstrates that with appropriate post-filtering, WGS can provide a scalable and clinically manageable CNV interpretation workflow.

### 3.5 Screening of FragX from WGS

In addition to evaluating the feasibility of replacing CMA with WGS, we also assessed whether WGS could replace repeat primed PCR for FMR1 (Fragile X) screening when ordered in conjunction with CMA. While repeat primed PCR is a low-cost and widely available assay, our cWGS platform uses short-read sequencing, which presents challenges in accurately sizing large trinucleotide repeats. Therefore, the goal of this assessment was not to replace PCR in all cases, but rather to determine whether WGS could reliably rule out Fragile X in negative cases.

In clinical production, WGS could serve as an initial screening tool, flagging potential positive cases for follow-up PCR confirmation to obtain precise repeat sizing.

To evaluate this approach, we analyzed 39 samples with known FMR1 repeat sizes from repeat primed PCR. To ensure high-confidence repeat sizing and prevent false negatives, we required each sample to have at least two reads fully spanning the CGG repeat region per allele in order to be considered confidently classifiable by WGS.

Table 3 summarizes the results after applying this high-confidence threshold on Expansion Hunter output. All 16 normal samples were correctly identified by WGS, with repeat lengths matching PCR results within +/-1 repeat, sufficient for clinical negative classification, One gray zone sample also met the high-confidence threshold and was correctly classified, though PCR confirmation would still be required. Importantly, all negative/normal samples were correctly identified, which represents the majority of Fragile X testing ordered alongside CMA.

**Table 3.** Summary of suboptimal matches between CMA and WGS.

| Event Type | Event Size | Reciprocal Overlap (%) | Reasons for Suboptimal Matches | More Accurate Platform |
| --- | --- | --- | --- | --- |
| DEL | 1.9Mb | 47.12 | KMER-NON-UNIQUE region is too wide | NA |
|  | 312kb | 66.11 | WGS has better resolution than CMA | WGS |
| DUP | 1.1Mb | 68.26 | WGS has better resolution than CMA | WGS |
| LOH | 15.8Mb | 65.51 | WGS has better resolution than CMA | WGS |
|  | 7Mb | 56.19 | WGS has better resolution than CMA | WGS |

**Table 4.** Summary of cWGS and PCR concordance for FMR1 repeat classification.

| Status | Repeat Size | Samples | cWGS High Confident REPCN* | Requires Confirmation |
| --- | --- | --- | --- | --- |
| Normal | < 40 | 16 | 16 | 0 |
| Gray Zone | 41-54 | 12 | 1 | 12 |
| Premutation | 55-199 | 4 | 0 | 4 |
| Full Expansion | > 200 | 7 | 0 | 7 |
| Total |  | 39 | 17 (43%) | 23 (58%) |

\* REPCN supported by at least 2 spanning reads.

These findings demonstrate that short-read WGS can achieve 100% sensitivity for ruling out Fragile X syndrome in negative cases, supporting its use case as a reliable initial screening tool to confidently exclude negatives while driving potential positives for confirmatory PCR testing.

## 4. Discussion

This study demonstrates that WGS can reliably detect clinically relevant CNVs and LOH across a broad range of sizes, achieving high concordance with CMA and supporting its use as a first-tier diagnostic test for pediatric patients with developmental disabilities and congenital anomalies. In addition, WGS enables efficient Fragile X screening and the consolidation of multiple traditional assays into a single workflow.

CMA has been recommended as a first-tier test for individuals with intellectual development disorders and congenital malformations since the 2010 consensus statement by ACMG^4^.

Following this guidance, CMA became widely adopted in clinical practice and is currently offered in many diagnostic laboratories ranging from large commercial reference labs to in-house Laboratory Medicine departments in medical centers or hospitals. While CMA is an excellent diagnostic tool, the data generated for CMA can only be used for this single assay. Any additional testing, like FragileX analysis or sequencing based tests like panels, WES, or WGS require additional laboratory work. However, with the continued reduction in sequencing costs and the comprehensive nature of WGS, there is increasing interest in adopting WGS as a first-line diagnostic tool^16,17,18,19^. Unlike CMA, WGS can act as a single laboratory assay that enables multiple analyses or clinical tests to be offered using the same data. These analyses can be performed serially or through cascade approaches without performing additional laboratory work. This can reduce cost to the testing laboratory and the patient and can significantly reduce the time to diagnosis.

A 97.28% concordance rate (95%CI: 93.80-98.83) between CMA and WGS underscores WGS’s robustness for CNV and LOH detection. In contrast, the detection of CNVs and LOH by CMA is inherently restrained by the density and distribution of probes across the genome. As a result, clinically significant CNVs may be missed, inaccurately sized, or misinterpreted. Specifically, imprecise breakpoint resolution can lead to incorrect interpretation of which gene or regulatory elements are affected, potentially obscuring important clinical findings. Among all suboptimal matches (N = 5) between CMA and WGS in CMA-derived solves, 80% of the discrepancies were driven by WGS providing more precise breakpoint resolution. This advantage stems from WGS’s broader and more uniform CNV data point coverage across the genome. Beyond individual case analyses, we systematically quantified the CNV data coverage by the two platforms. WGS offers coverage of 83.55% genome-wide and 97.38% in clinical relevant regions, compared to less than 3% with CMA. Together, these results collectively highlight the superiority of WGS over CMA for CNV detection.

The increased resolution of WGS also results in a substantially higher number of CNV calls which requires manual clinical interpretation. Previous studies have shown WGS can generate thousands of SVs calls per sample, depending on the calling algorithm and filtering strategy^20,21,22^. While many of those events are biologically informative and provide valuable insights for population genetics, they are not all suitable for clinical diagnostic setting. To address this challenge, prior studies have applied population frequency and genomic context annotation to help distinguish rare, potentially pathogenic events from common, likely benign ones in WGS-derived CNV calls^14,21^. Building on these approaches, we implemented a cohort-based occurrence filter tailored to our sample set and CNV calling pipeline. This strategy effectively prioritizes rare and potentially pathogenic CNVs, significantly reducing the interpretive burden without compromising clinical sensitivity, as none of the reported CNV solves were lost post-filtering.

To support Fragile X screening, which is often ordered together with the CMA test, we developed a custom PCR confirmation logic based on Expansion Hunter output. By leveraging the number of spanning reads and the confidence interval of repeat estimate, our logic allows WGS to confidently exclude cases with fewer than 35 FMR1 repeats and achieve +/- 1 repeat concordance with PCR results. Ambiguous or potentially expanded alleles are correctly flagged for PCR confirmation.

This study was designed to assess whether WGS can match the sensitivity of CMA for detecting clinically relevant CNVs and LOH, rather than evaluating its potential for increased diagnostic yield. As such, our analysis emphasized on concordance and clinical feasibility rather than novel discoveries. However, prior studies have demonstrated that WGS can uncover additional clinically relevant CNVs not detected by CMA, particularly in regions with strong bias in probe placement^6,16,20^. Moreover, WGS provides the opportunity to identify incidental findings with potential clinical relevance, further enhancing its diagnostic value^20,24,25^. Further studies are needed to systematically evaluate the added diagnostic benefits of WGS compared to CMA.

Together, our results demonstrate that WGS, when paired with appropriate filtering and logic, can serve as a scalable and effective alternative to CMA in clinical practice. As sequencing costs continue to fall and analytical tools improve, our results provide a strong case for adopting WGS as the new standard first-tier test in clinical testing, enabling faster, more accurate, and more efficient diagnostic experience for patients with complex genetic disorders.

## Data Availability

All data produced in the present study are available upon reasonable request to the authors.

## Declaration of AI and AI-assisted technologies in the writing process

During the preparation of this work, the authors used ChatGPT to assist with editing for clarity and conciseness. After using this tool, the authors reviewed and edited the content as needed and take full responsibility for the final version of the manuscript.

## Acknowledgement

We thank the DRAGEN SV team, including Sean Truong, Francesco Brundu, Yeonghun Lee for their valuable feedback and support throughout this project. Their input helped with optimizing the WGS-based CNV and LOH detection in the DRAGEN cytogenetics module.

## Author contribution

Conceptualization: SS, MF, KL, SH, NL, PB

Data Curation: YG, SH, PB, CC, JLC, BAH, GL, SWM, RMT

Formal Analysis, YG, SH

Investigation: YG, SS, MF, KL, VP, SH, NL, PB

Methodology: YG, VP, SH, SS

Visualization: YG, SH, PB

Writing – original draft: YG, SS, KL, SH

Writing – review & editing: YG, SS, CC, JLC, MF, BAH, KL, GL, SWM, VP, RMT, SH, NL, PB

## Disclosure

N.L. received personal fees from Illumina Inc and is a scientific advisory board member for FYR Diagnostics, ML4H, and Everygene, Inc. V.P. is a former employee and owns shares of Illumina.

## Notes

### Funding Statement

This study was funded by Quest Diagnostics and Broad Clinical Labs

### Author Declarations

Institutional Review Board of WCG gave ethical approval for this work (IRB protocol identifier 20242498).

