## Supplemental Notes for "Whole-Genome Sequencing is a Viable Replacement for Chromosomal Microarray and Fragile X PCR Testing"

### Supplemental Note 1: Different representation by two platforms but essentially the same call

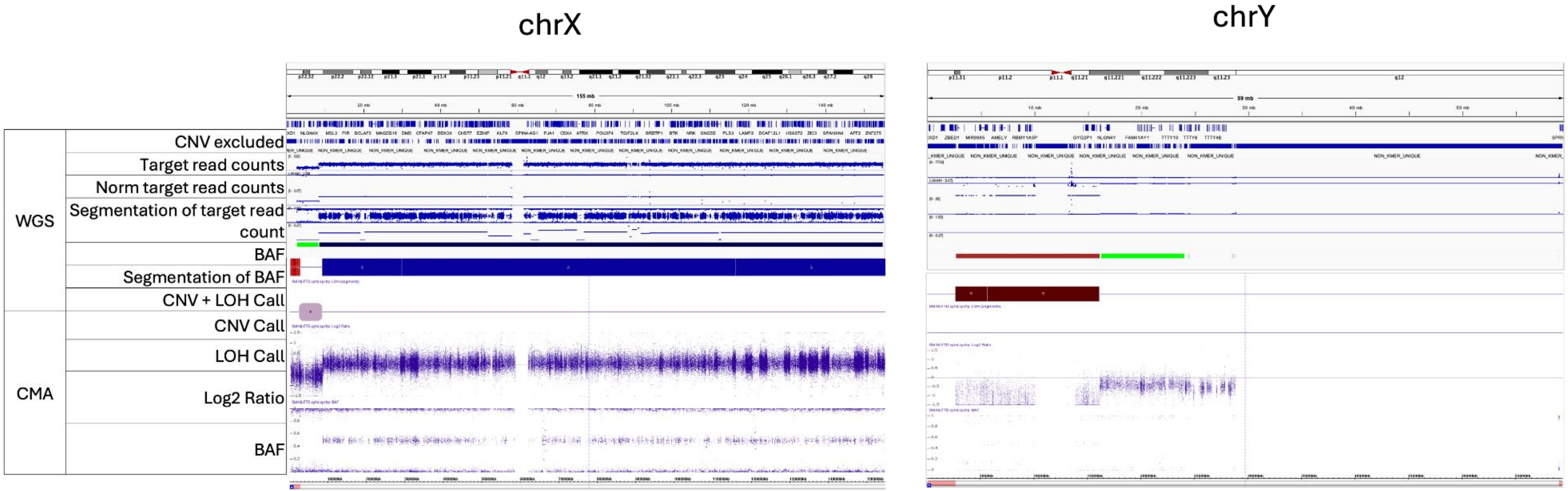


CMA analysis identified complex sex chromosome abnormalities in one sample, initially missed by WGS's automated pipeline. The array classified the sample as a XY individual  and reported the following: arr[GRCh37] Xp22.33p22.31(168547_8428908)x1, Yq11.221q11.23(16099876_28799937)x1. Upon manual review, WGS data showed signals consistent with these findings but presented the regions differently, resulting in initial omission by automated evaluation process.

Supplemental Note 2: Sub calls were manually merged by Quest during clinical interpretation

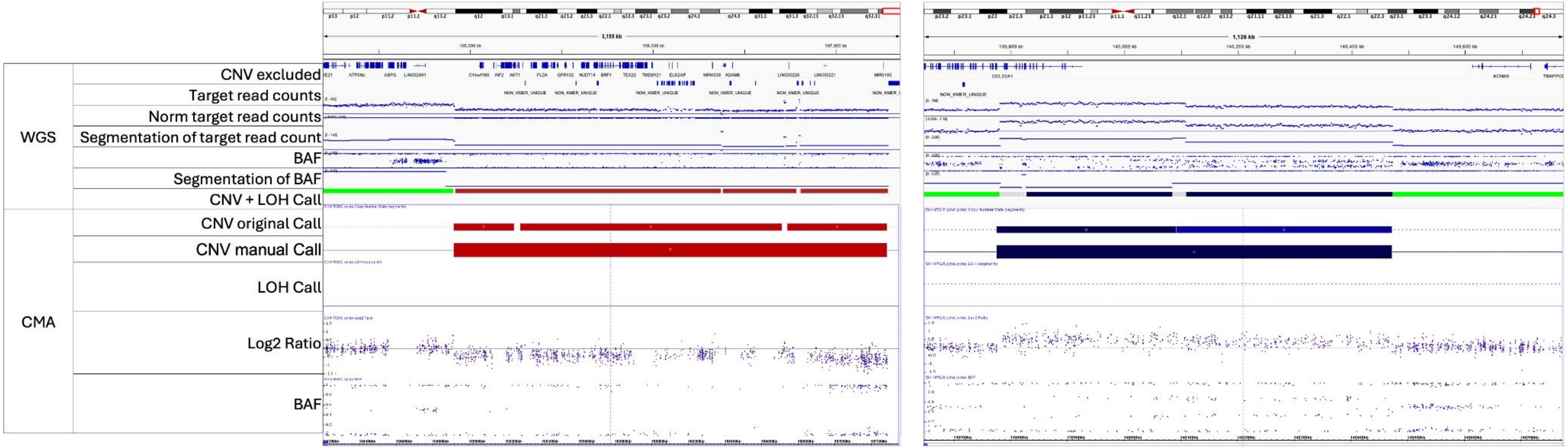
